# Bayesian joint modelling framework linking antibody kinetics and test-negative vaccine effectiveness to study hybrid immunity across epidemic waves

**DOI:** 10.64898/2026.04.25.26351732

**Authors:** Aimane Benammar

## Abstract

Vaccine effectiveness against symptomatic SARS-CoV-2 infection varies over time and across epidemic waves. This variation can reflect waning immunity, immune escape by emerging variants, exposure heterogeneity, and differences in previous infection history. Test-negative case-control designs are widely used to monitor vaccine effectiveness, while longitudinal serological studies describe antibody trajectories after vaccination and infection. These evidence streams are often analysed separately.

This manuscript presents a simulation-based Bayesian joint modelling framework that links individual-level antibody kinetics to test-negative vaccine effectiveness estimates across successive epidemic waves. Hybrid immunity is represented as the combined effect of vaccination and infection history, with latent antibody titres following a boost-and-decay process after each immunising event. A variant-specific titre-protection curve maps latent antibody levels to the risk of symptomatic infection. The framework is intended to illustrate how apparent changes in vaccine effectiveness may be decomposed into components related to waning, immune escape, and exposure heterogeneity.

Using fully synthetic data calibrated to plausible vaccination schedules, infection histories, assay variability, and epidemic-wave structures, the model is evaluated in three simulation studies. The simulations illustrate that joint modelling can recover broad features of the assumed titre-protection relationship under idealised conditions and can separate waning from variant-specific shifts when the data-generating process is correctly specified. The results are not presented as validation on real-world surveillance data. Instead, they provide a transparent methodological proof of concept and identify assumptions that would need to be assessed before applying the framework to linked serological and test-negative datasets.

**Author declarations:** This manuscript reports a methodological simulation study. All individual-level data used in the manuscript are synthetic. No human participants, patient records, biological samples, or identifiable data were used. No ethics approval was required for the analyses presented here. The author declares no competing interests. This study did not receive external funding.

## 1 Introduction

Test-negative designs have become a standard observational approach for estimating vaccine effectiveness against respiratory infections, including COVID-19 [3, 5, 8]. In these studies, symptomatic individuals who seek testing are classified as test-positive cases or test-negative controls, and vaccine effectiveness is estimated from the association between vaccination status and test positivity. The design is useful because it can reduce some forms of healthcare-seeking bias, but interpretation remains challenging when vaccination, previous infection, exposure risk, and variant circulation change over time.

Neutralising antibody levels have been shown to correlate with protection against symptomatic SARS-CoV-2 infection across vaccine platforms and studies [4, 6]. At the same time, antibody concentrations decline after immunising events, while emerging variants may require higher neutralisation levels to achieve similar protection. These mechanisms make vaccine-effectiveness trajectories difficult to interpret when analyses are performed only at the population level.

Hybrid immunity, here defined as immunity arising from both vaccination and infection, adds another layer of complexity. Empirical studies and reviews have reported that combined exposure to vaccination and infection can produce broader or stronger immune responses than either exposure alone, although the magnitude and durability of protection vary by variant, population, assay, and time since exposure [1, 2, 7]. A modelling framework that jointly represents serological dynamics and test-negative outcomes may therefore help clarify which part of an observed vaccine-effectiveness decline is compatible with waning immunity and which part is compatible with immune escape or exposure changes.

The objective of this manuscript is not to estimate vaccine effectiveness from real surveillance data. Instead, the aim is to describe and stress-test a Bayesian joint modelling framework using synthetic data only. The contribution is methodological. The manuscript shows how the model can be specified, what quantities can be estimated under controlled simulation conditions, and which assumptions require validation before any real-world application.

## 2 Conceptual framework

Figure 1 summarises the framework. Immunising events generate latent antibody trajectories. These trajectories are mapped to variant-specific protection through titre-protection curves. Protection, exposure intensity, and covariates determine infection risk, while symptomatic testing creates the synthetic test-negative sample.

**Figure 1:**
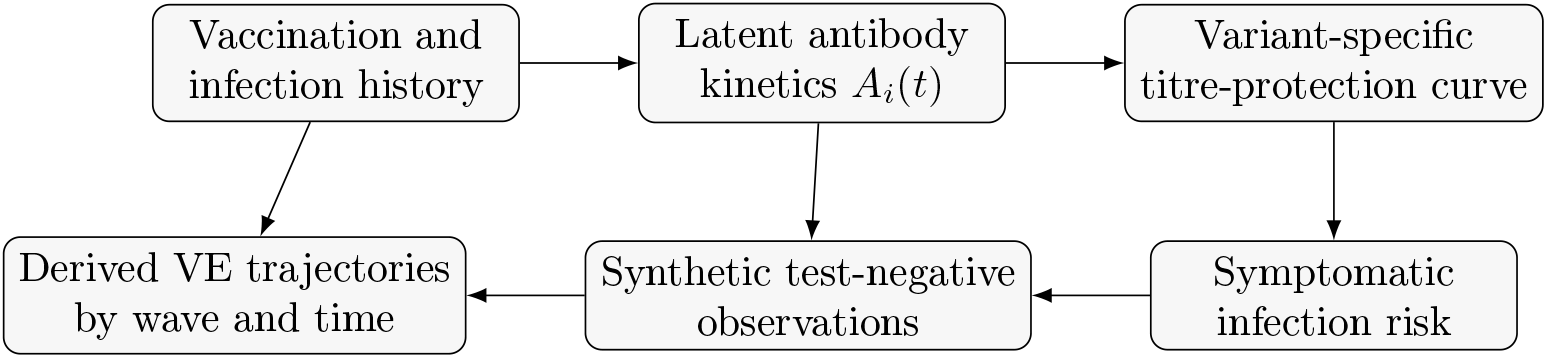
Conceptual structure of the simulation-based joint model. The figure is schematic and does not represent real participant data.

## 3 Methods

### 3.1 Synthetic cohort and epidemic waves

A synthetic cohort is generated with vaccination times, infection histories, covariates, and testing times. The simulation contains three stylised epidemic waves: an ancestral-like wave, an intermediate immune-escape wave, and a stronger immune-escape wave. These waves are not intended to reproduce any specific country, period, or variant lineage.

Each simulated individual belongs to one of three simplified immunity groups: vaccine-only, infection-only, or hybrid immunity. These categories are used only for model illustration. In practice, real-world immunity is more granular and depends on vaccine product, dose number, infection timing, variant exposure, immune status, age, and other factors.

### 3.2 Antibody kinetics model

Let *A*_*i*_(*t*) denote the latent log antibody titre of individual *i* at time *t*. The synthetic data-generating process assumes that each immunising event *k* produces a boost followed by exponential decay:

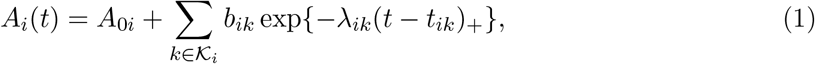

where *A*_0*i*_ is a baseline term, *t*_*ik*_ is the time of immunising event *k, b*_*ik*_ is the event-specific boost, *λ*_*ik*_ is the waning rate, and (*x*)_+_ = max(*x*, 0). In the Bayesian model, event-specific boosts and waning rates are given hierarchical priors:

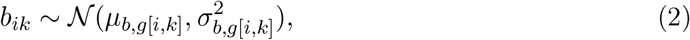

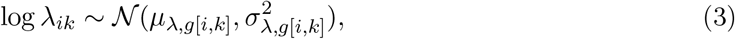

where *g*[*i, k*] indexes the exposure class. Observed antibody measurements, when simulated, are generated with assay noise:

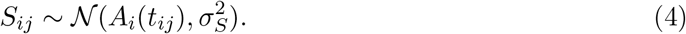

### 3.3 Titre-protection relationship

For wave or variant scenario *v*, the probability of symptomatic infection for individual *i* at time *t* is modelled as:

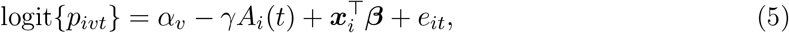

where *α*_*v*_ is a wave-specific baseline risk parameter, *γ >* 0 controls the association between antibody titre and protection, ***x***_*i*_ are covariates, and *e*_*it*_ represents exposure intensity. Immune escape is represented by a shift in *α*_*v*_ or, equivalently, by a higher titre threshold required to achieve the same level of protection. This formulation is a simplified statistical representation inspired by correlate-of-protection work [4, 6]; it is not a complete immunological model.

### 3.4 Synthetic test-negative component

The test-negative sample is generated by simulating symptomatic testing events. For each tested individual, a binary test outcome *Y*_*i*_ is generated, where *Y*_*i*_ = 1 indicates test-positive symptomatic infection and *Y*_*i*_ = 0 indicates a test-negative symptomatic control. Conditional on inclusion in the synthetic test-negative sample, the model links *Y*_*i*_ to the latent antibody titre at test time, wave membership, exposure intensity, vaccination status, and covariates.

Vaccine effectiveness is derived as a model-based contrast between predicted infection risks under observed vaccination history and a counterfactual vaccination history, while keeping specified covariates fixed. For a target group *G* at time *t* in wave *v*:

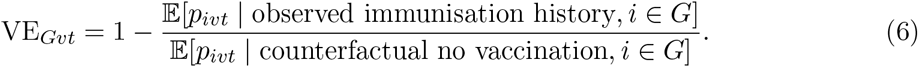

This quantity is a model-based estimand in the synthetic system, not an empirical estimate from real surveillance data.

### 3.5 Bayesian inference and simulation scenarios

Posterior inference can be performed using Markov chain Monte Carlo or variational inference. The posterior distribution combines the antibody observation model and the synthetic test-negative likelihood. Weakly informative priors are used for regression coefficients and hierarchical variance parameters.

The manuscript evaluates three qualitative simulation studies. The first examines antibody kinetics under different immunity scenarios. The second examines vaccine-effectiveness trajectories across stylised epidemic waves. The third examines a counterfactual decomposition of apparent vaccine-effectiveness loss into components compatible with waning and immune escape.

## 4 Results

### 4.1 Overview of the synthetic experiments

The results presented in this section are based entirely on synthetic data generated from the assumptions described in the Methods section. The purpose is not to estimate vaccine effectiveness in any real population, but to examine whether the proposed Bayesian joint modelling framework behaves coherently under controlled simulation conditions.

Three synthetic experiments were considered. The first experiment examined antibody titre trajectories after simplified immunising histories. The second experiment translated these titre trajectories into stylised vaccine-effectiveness curves across three epidemic-wave scenarios. The third experiment illustrated a counterfactual decomposition of apparent vaccine-effectiveness loss into components compatible with waning immunity and immune escape.

The numerical values used in the figures are arbitrary and were chosen only to create interpretable synthetic scenarios. They should therefore be understood as model-generated examples rather than empirical estimates.

### 4.2 Synthetic antibody kinetics

Figure 2 shows illustrative geometric mean titre trajectories after the most recent immunising event under three stylised synthetic scenarios. Under the assumed simulation parameters, the hybrid-immunity scenario generates a higher initial titre and a slower apparent decline than the vaccine-only and infection-only scenarios.

**Figure 2:**
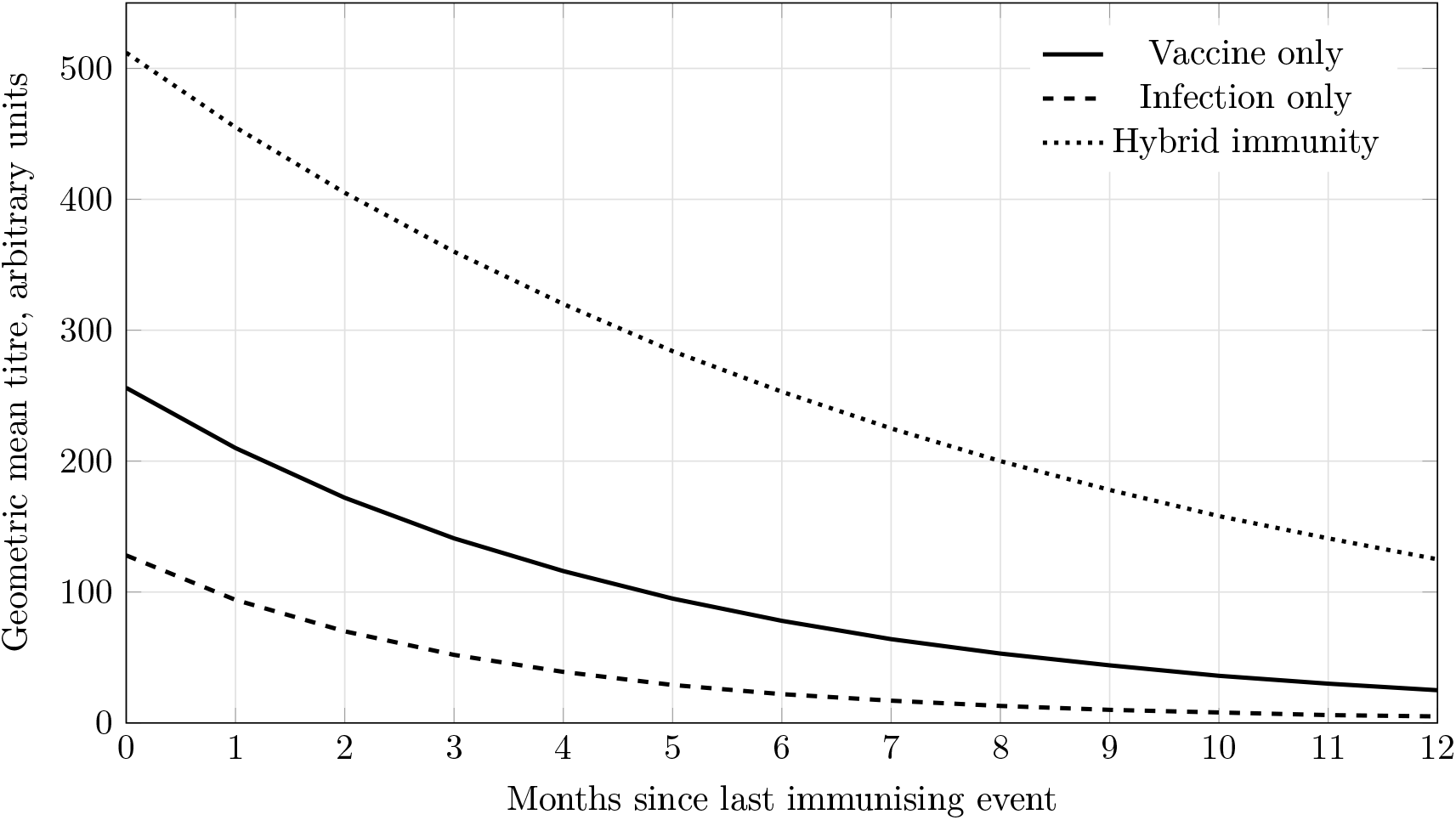
Illustrative synthetic geometric mean titre trajectories under three stylised immunising histories. Values are arbitrary and were generated solely for model demonstration.

This pattern is expected because the data-generating process explicitly assigns stronger boosting and slower waning to the hybrid-immunity scenario. The figure should therefore be interpreted as an internal check that the simulated antibody module reflects the intended qualitative behaviour, rather than as evidence about real antibody kinetics.

### 4.3 Synthetic vaccine-effectiveness trajectories across waves

Figure 3 shows model-derived vaccine-effectiveness trajectories under three stylised wave scenarios. The ancestral-like scenario has the highest initial protection, whereas immune-escape scenarios have lower apparent protection because the assumed titre-protection curve is shifted. The decline over time reflects antibody waning under the assumed kinetics. In this synthetic setting, the difference between waves is not caused by changes in the simulated population alone, but by the assumed variant-specific shift in the relationship between antibody titre and protection.

**Figure 3:**
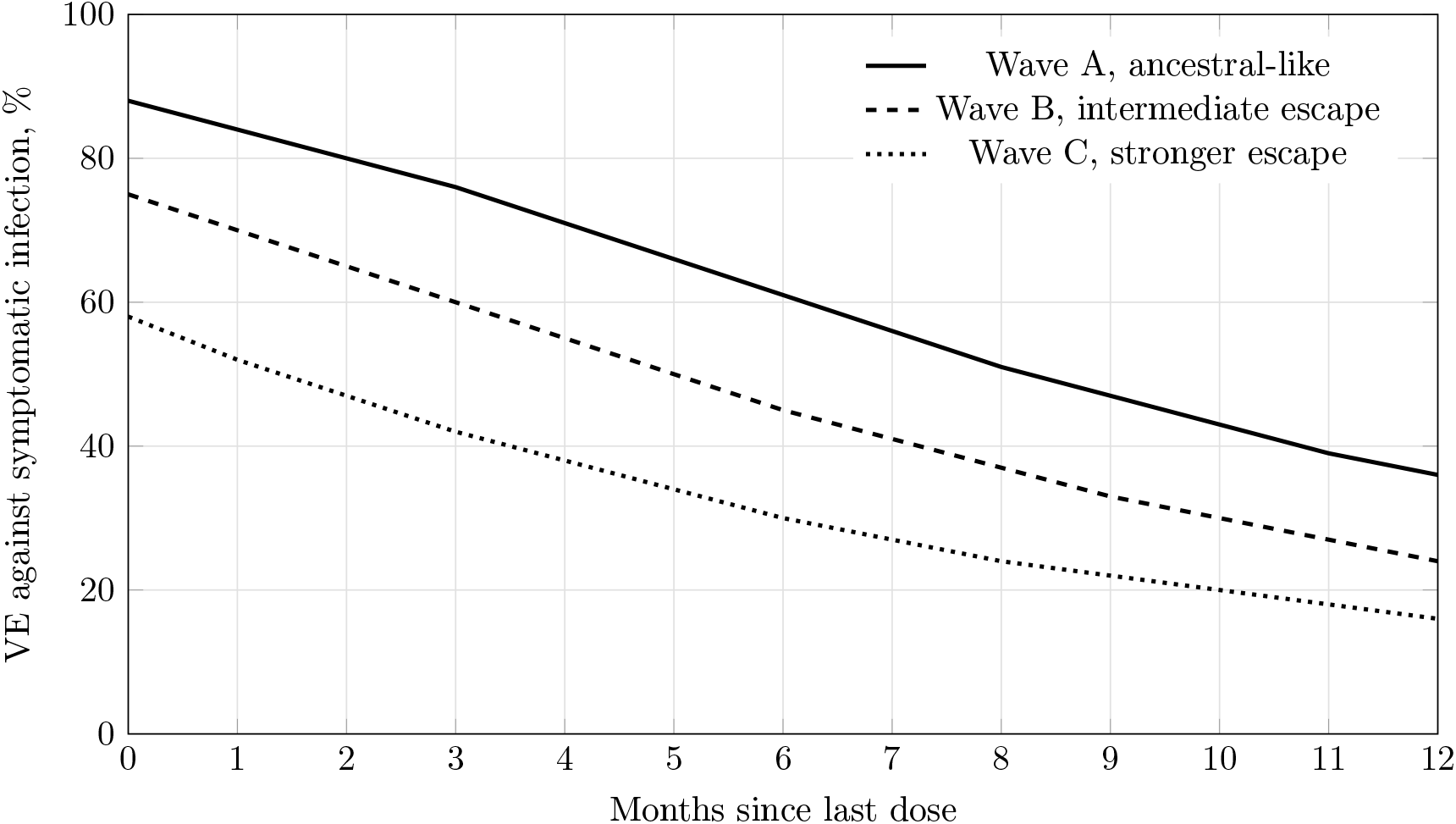
Illustrative synthetic vaccine-effectiveness trajectories across three stylised wave scenarios. These curves were generated from the assumed simulation model and do not represent estimates from real surveillance data.

### 4.4 Counterfactual decomposition of apparent vaccine-effectiveness loss

Figure 4 illustrates a counterfactual decomposition for the stronger immune-escape scenario. The synthetic observed trajectory combines a lower initial protection level with subsequent waning. The counterfactual trajectory is generated under the same waning structure but without the immune-escape shift in the titre-protection curve.

**Figure 4:**
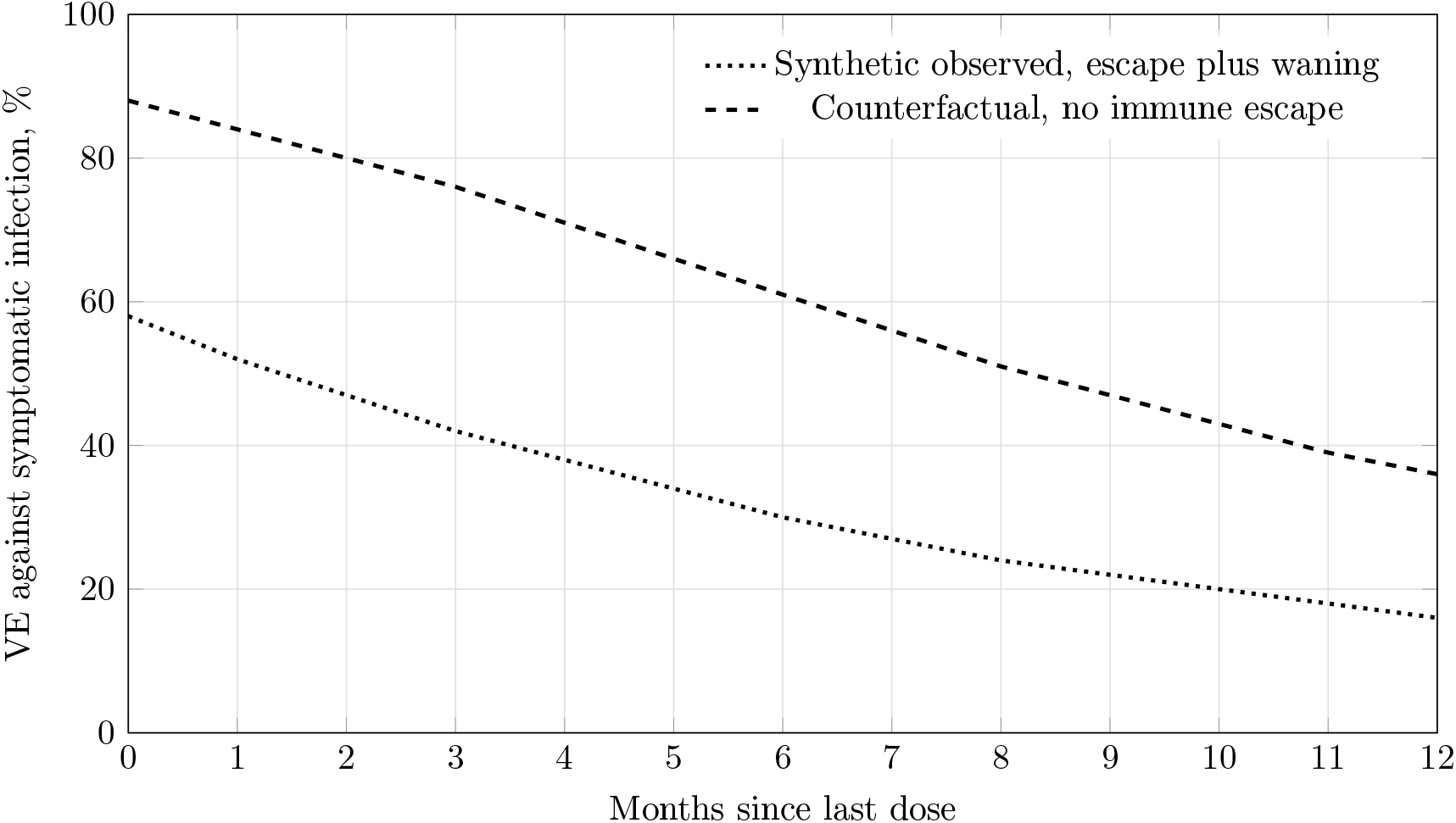
Illustrative counterfactual decomposition of apparent vaccine-effectiveness loss in a synthetic immune-escape wave. The decomposition is conditional on the assumed data-generating model and should not be interpreted as a real-world estimate.

Within the simulation, the gap between the two curves is interpreted as the component compatible with immune escape. This interpretation is conditional on the assumed data-generating process. In real-world data, such a decomposition would require stronger assumptions and careful sensitivity analyses because waning, exposure, prior infection, testing behaviour, and variant circulation may be correlated.

### 4.5 Interpretation of the simulation outputs

Across the three synthetic experiments, the framework behaves as expected under the assumptions used to generate the data. Higher simulated antibody titres are associated with higher predicted protection, immune-escape waves shift the protection curve downward, and waning produces a gradual decline in model-derived vaccine effectiveness over time.

These findings should not be interpreted as evidence that the same decomposition would be identifiable in real data. Rather, they show that the proposed joint model provides a coherent structure for connecting immunological dynamics and test-negative outcomes. The main methodological advantage is that the model makes assumptions explicit. This allows researchers to examine how changes in antibody kinetics, exposure intensity, or variant-specific protection thresholds affect vaccine-effectiveness trajectories.

The results also highlight the importance of linked data. In real applications, the ability to distinguish waning immunity from immune escape would likely depend on the availability of vaccination dates, infection history, repeated serological measurements, variant or wave information, and covariates related to exposure and testing behaviour.

## 5 Model checking and sensitivity analysis

Because this manuscript uses synthetic data, model checking focuses on internal consistency rather than empirical validation. In a complete implementation, posterior predictive checks would compare simulated test-positive proportions, antibody titre distributions, and vaccine-effectiveness trajectories with the corresponding quantities generated by the model.

Several sensitivity analyses would be important before applying the framework to real-world data. First, the antibody waning function could be varied from exponential decay to biphasic or spline-based decay. Second, the titre-protection relationship could be specified using alternative functional forms, including logistic, probit, or threshold-based curves. Third, exposure heterogeneity could be modelled using fixed covariates, random effects, or time-varying intensity terms.

Fourth, undocumented infections could be introduced as latent events, with prior distributions informed by seroprevalence or external surveillance.

These sensitivity analyses are important because the decomposition of apparent vaccine-effectiveness loss into waning and immune escape is not automatically identifiable. If variant circulation, time since vaccination, previous infection, and exposure risk are strongly correlated, different mechanisms may generate similar observed vaccine-effectiveness trajectories. The framework is therefore best viewed as a structured tool for assumption-based inference rather than as a direct estimator that can separate biological mechanisms without additional information.

## 6 Discussion

This simulation study presents a Bayesian framework for jointly modelling antibody kinetics and test-negative vaccine-effectiveness outcomes. The primary value of the framework is interpretability. Rather than treating vaccine-effectiveness trajectories as isolated regression outputs, the model expresses them through latent antibody dynamics, variant-specific shifts in the titre-protection relationship, and synthetic testing outcomes generated under an explicit set of assumptions.

Under idealised assumptions and correct model specification, the simulations recover qualitative patterns that were built into the data-generating process. The framework also produces derived quantities that separate apparent vaccine-effectiveness loss into components compatible with waning and immune escape. This is useful as a methodological proof of concept, but it is not sufficient to establish real-world performance, causal validity, or policy relevance.

The proposed framework has several potential strengths. First, it creates a direct link between immunological measurements and population-level vaccine-effectiveness analyses. Second, it allows uncertainty to be propagated through antibody kinetics, titre-protection curves, and test-negative outcomes in a single Bayesian model. Third, it encourages transparent assumptions about immune escape and waning, rather than attributing all temporal changes in vaccine effectiveness to a single mechanism.

Several limitations should be emphasised. First, all data used in this study are synthetic. The reported figures should therefore not be interpreted as empirical estimates for any country, population, vaccine product, or variant lineage. Second, the antibody model is deliberately simplified and does not account for cellular immunity, mucosal immunity, immune imprinting, assay-specific measurement error, or broader sources of immunological heterogeneity. Third, test-negative designs may be affected by changing healthcare-seeking behaviour, differential testing access, exposure heterogeneity, outcome misclassification, and undocumented infections, none of which are fully represented in the present simulations. Fourth, the identifiability of waning and immune escape may be weak unless serology, vaccination history, infection history, epidemic timing, and relevant covariates are jointly informative.

Future work should assess this framework using appropriately governed real-world linked datasets only when the necessary ethical approvals, data-access permissions, and privacy safeguards are in place. Such work should include posterior predictive checks, sensitivity analyses for exposure heterogeneity and undocumented infection, comparisons with conventional test-negative analyses, and transparent reporting of model assumptions and limitations.

## 7 Conclusion

A Bayesian joint modelling framework can provide a coherent way to connect synthetic antibody kinetics and synthetic test-negative outcomes in order to study how waning immunity and immune escape may shape vaccine-effectiveness trajectories. The present manuscript should be read as a simulation-based methodological proof of concept. It makes no claim of real-world vaccine-effectiveness estimation and requires validation on governed real-world datasets before operational or policy use.

## Data availability

All data used in this manuscript are synthetic. The simulation design and model equations are described in the manuscript. No individual-level human participant data were used. Simulation code and analysis scripts should be deposited in a public repository before or at the time of preprint posting to support reproducibility.

## Ethics statement

This manuscript reports a methodological simulation study using synthetic data only. It did not involve human participants, patient records, biological samples, or identifiable data. Ethics approval was therefore not required.

